# Multi-task Deep Learning Based CT Imaging Analysis For COVID-19: Classification and Segmentation

**DOI:** 10.1101/2020.04.16.20064709

**Authors:** A. Amyar, R. Modzelewski, S. Ruan

## Abstract

The fast spreading of the novel coronavirus COVID-19 has aroused worldwide interest and concern, and caused more than one million and a half confirmed cases to date. To combat this spread, medical imaging such as computed tomography (CT) images can be used for diagnostic. An automatic detection tools is necessary for helping screening COVID-19 pneumonia using chest CT imaging. In this work, we propose a multitask deep learning model to jointly identify COVID-19 patient and segment COVID-19 lesion from chest CT images. Our motivation is to leverage useful information contained in multiple related tasks to help improve both segmentation and classification performances. Our architecture is composed by an encoder and two decoders for reconstruction and segmentation, and a multi-layer perceptron for classification. The proposed model is evaluated and compared with other image segmentation and classification techniques using a dataset of 1044 patients including 449 patients with COVID-19, 100 normal ones, 98 with lung cancer and 397 of different kinds of pathology. The obtained results show very encouraging performance of our method with a dice coefficient higher than 0.78 for the segmentation and an area under the ROC curve higher than 93% for the classification.

## 1. INTRODUCTION

The novel coronavirus disease (COVID-19) spread rapidly around the world, changing the daily lives of billions of people. The infection can lead to severe pneumonia that can causes death. Also, COVID-19 is highly contagious, which is why it must be detected quickly, in order to isolate the infected person very fast to limit the spread of the disease.Today, the gold standard for detecting COVID-19 is the Reverse Transcription Polymerase Chain Reaction (RT-PCR) [1], which consists of detecting viral RNA from sputum or nasopharyngeal swab. The limitation with the RT-PCR test is due to the time needed to get the results, the availability of the material which remains very limited in hospitals [1] and its relatively low sensitivity, which does not meet the major interest of rapidly detecting positive cases as soon as possible in order to isolate them [2]. An alternative solution for rapid screening could be the use of medical imaging such as x-chest ray images or computed tomography (CT) scanners [2].

Identifying COVID-19 at an early stage through imaging would indeed allow the isolation of the patient and therefore limit the spread of the disease [2]. However, physicians are very busy fighting this disease, hence the need to create decision support tools based on artificial intelligence to not only detect but also segment the infection at the lung level in the image [1]. Artificial intelligence has seen a major and rapid growth in recent years with deep neural networks [3] as a first tool to solve different problems such as object detection [4], speech recognition [4], and image classification [5]. More specifically, convolutional neural networks (CNNs) [6] showed astonishing results for image processing [7]. For image segmentation, several works have shown the power and robustness of these methods [8]. CNNs architectures for medical imaging also have been used with very good results [9], for both image classification [10] or image segmentation [11].

### 1.1. Related Work

For the detection of COVID-19 and the segmentation of the infection at the lung level, several deep learning works on x-chest ray images and CT scans have emerged and reported in [12]. In [13] Ali Narin et al. created deep convolutional neural networks to automatically detect COVID-19 on X-ray images. To that end, they used transfer learning based approach with a very deep architectures such as ResNet50, InceptionV3 and Inception-ResNetV2. The algorithms were trained on the basis of 100 images (50 COVID vs 50 non-COVID) in 5 cross-validation. Authors claimed 97 % of accuracy using InceptionV3 and 87% using Inception-ResNetV2, however, due to the very limited size of patient and the very deep models, overfiting would rise and could not be ruled-out, hence the need to validate those results in a larger database is necessary. Also in [14], Hemdan et al. created several deep learning models to classify x-ray images into COVID vs non-COVID classes reporting best results with an accuracy of 90% using VGG16. Again, the database was very limited with only 50 cases (25 COVID vs 25 non-COVID). A resembling study was conducted by wang and wang [15] where they trained a CNN on the ImageNET database [16] then fine-tuned on x-ray images to classify cases into one of four classes: normal, bacterial, non-COVID-19 viral and COVID-19 viral infection, with an overall performance of 83.5%. For CT images, Jinyu Zhao et al [17] created a container for CT scans initially with 275 CT COVID-19 on which they also applied a transfer learning algorithm using chest-x-ray14 [18] with 169-layer DenseNet [19]. The performance of the model is 84.7% with an area under the ROC curve of 82.4%. As of today, the database contains 347 CT images for COVID-19 patients and 397 for non-COVID patients.

Instead of using CNNs, other works have used network capsules which were first proposed in [20] to solve the problems of lack of data, and the needs for CNNs of data-intensive and many parameters. In the study [21] where the authors opted for this method, they created a capsule network to identify COVID-19 cases in x-ray images. The results were encouraging with an accuracy of 95.7%, sensitivity at 90% and specificity at 95.8%. They compared their results with Sethy et al. [22] where they created a model based on resnets50 with SVMs and obtained a performance of 95.38%, a sensitivity of 97.29% and a specificity of 93.47%.

In [23], Jin et al. created and deployed an AI tool to analyze CT images of COVID-19 in 4 weeks. To do this, a multidisciplinary team of 30 people collaborated together using a database of 1136 images including 723 positive COVID-19 images from five hospitals, to achieve a sensitivity of 0.974 and a specificity of 0.922. The system was deployed in 16 hospitals and performed over 1300 screenings per day. They proposed a combined model for classification and segmentation showing lesion regions in addition to the screening results. The pipeline is divided into 2 steps: segmentation and classification. They used several models including 3D U-NET++, V-NET, FCN-8S for segmentation and InceptionV3, ResNet50 and others for classification. They were able to achieve a dice coefficient of 0.754 using 3D U-NET ++ trained on 732 cases. The combination of 3D U-NET ++ and ResNet50 resulted in an area under the OCR curve of 0.991 with a sensitivity of 0.974 and a specificity of 0.922. In practice, the model continued to improve by re-training. The model proved to be very useful to physicians by highlighting regions of lesions which improved the diagnosis. What should be noted here is that the two models are independent and therefore they cannot help each other to improve both classification and segmentation performances.

### 1.2. Motivation

Multi-task learning (MTL) [24] is a type of learning algorithm whose goal is to combine several pieces of information from different tasks to improve the performance of the model and its ability to better generalize [25]. The basic idea of MLT is that different tasks can share a common features representation [25], and therefore, training them jointly. Using the whole dataset for the different tasks yield in a powerful representation and can improve performance for each task. Different approaches can be used in MTL such as hard parameter sharing [24] or soft parameter sharing [26]. Hard parameter sharing is the most commonly used approach to MTL in neural networks and greatly reduces the risk of overfitting [26]. It is generally applied by sharing the hidden layers between all tasks, while keeping several task-specific output layers. Soft parameter sharing defines a model for each task with its own parameters, and the distance between the parameters of the model is regularized in order to encourage the parameters to be similar.

In this work, we propose a novel multi-task deep learning model for jointly detecting COVID-19 image and segmenting lesions. The main challenges of this work are: 1) the lack of data and annotated data, the databases were collected from multiple sources with a huge variation in images and most of the images are not clean (see Fig 1), 2) instead of expensive models like ResNet 50 or DenseNet, developing a multitasking approach to reduce overfitting and improve results.

**Fig. 1.**
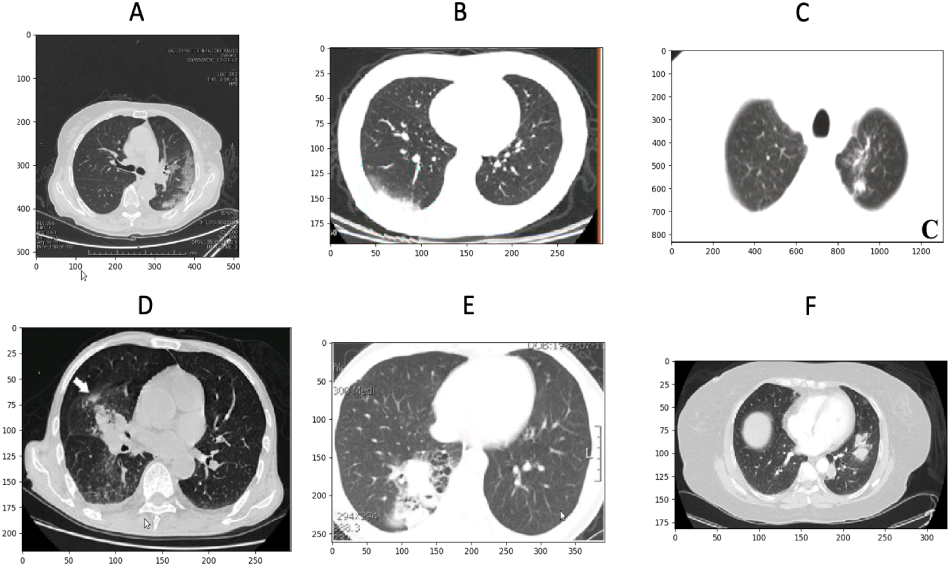
An example of different CT images for COVID (upper) and non-COVID (bottom) images. The similarities between certain COVID and non-COVID images make it difficult to generalize the model. In addition, patient images do not have the same resolution.Also, images show different image format (png(B C E F), Nifti (D), DICOM (A), different level of Visualisation Window (strong centering on lung (B E C), low (A,D, F), different images size in pixels (All), with/without annotation (with (C,D E), without (A B F), at different heights position in the lungs.

The paper is organized as follows. In Section 2, we describe our multi-task model, which is mainly based on classification and segmentation tasks. Section 3 presents the experimental studies. In section 4, we describe the validation methodology used in this study. Section 5 is showing the results of our work. Section 6 and 7 are for discussion and conclusion respectively.

## 2. METHOD

### 2.1. Data

In this study, three datasets including one thousand and forty four CT images are used.The first one is a public available data set coming from [17] which includes 347 COVID-19 images and 397 non-COVID images with different kinds of pathology. The database was pre-processed and stored in png format. The dimension varies from 153 to 1853 with an average of 491 for the height, while the width varies from 124 to 383 with an average of 1485 (see Fig 3). The second dataset coming from http://medicalsegmentation.com/covid19/ in which 100 COVID-19 CT scan with lesion ground truths are available. Three lesion labels are provided : ground glass, consolidation and plural effusion. As all legion labels are not given in all images, for the purpose of this study, we merged the three labels into one lesion label (See Fig 2). The third dataset coming from the hospital “Henri Becquerel Center” in Rouen city of France includes 100 CT of normal patients and 98 of lung cancer. All the three image datasets were resized to have the same size of 256 × 256 and the intensity normalized between 0 and 1 prior to analysis. Table 1 summarizes how to split the datasets for training, validation and test. All procedures performed in this study were conducted according to the principles expressed in the Declaration of Helsinki. The study was approved as a retrospective study by the Henri Becquerel Center Institutional Review Board. All patient information was de-identified and anonymized prior to analysis.

**Table 1.** Statistics of data split

|  | Non-COVID | COVID | Total |
| --- | --- | --- | --- |
| Train | 495 | 349 | 844 |
| Validation | 50 | 50 | 100 |
| Test | 50 | 50 | 100 |

**Fig. 2.**
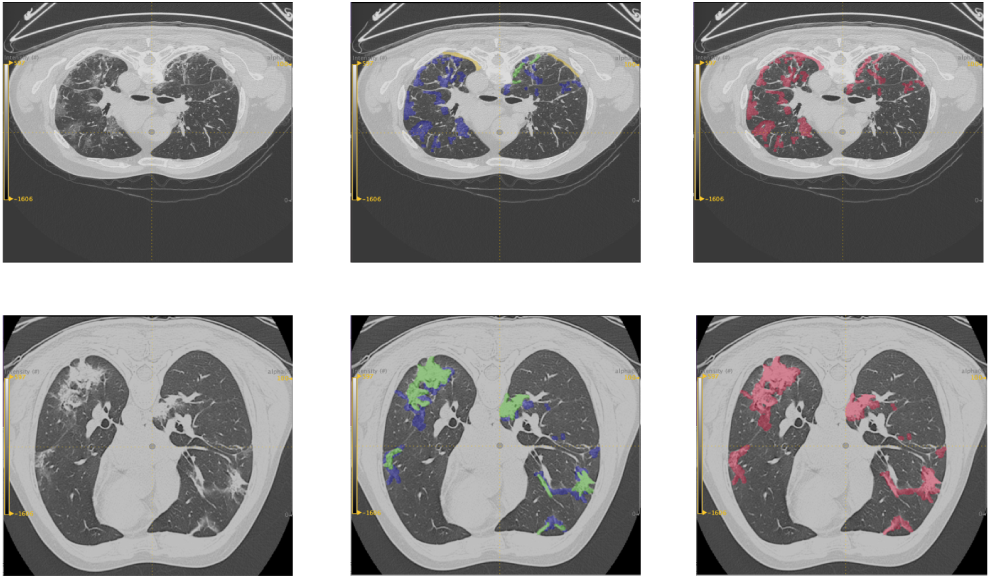
An example of the 3 labels: ground-glass (green), consolidation (blue) and pleural effusion (yellow) of two COVID-19 patients – 2D CT slice – 2D CT slice fused with 3 class segmentation - 2D CT slice fused with one class of pathology in red.

**Fig. 3.**
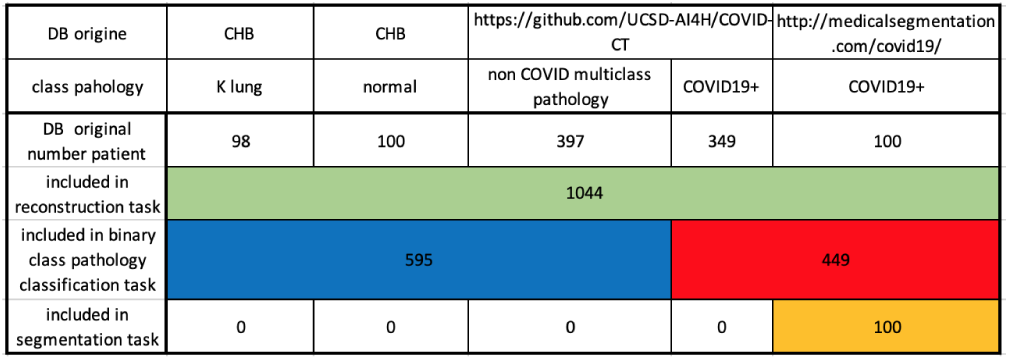
The different databases used in this study.

### 2.2. Model description

We have developed a new MTL architecture based on 3 tasks: 1) COVID vs non-COVID classification, 2) COVID lesion segmentation, 3) Image reconstruction. The two first tasks are essential ones, while the third task is added to enhance the feature representation extracted. In this work we choose to use a hard parameter sharing to share parameters between the different tasks. We create a common encoder for the three tasks which takes a CT scan as input, and its output is then used to the reconstruction of the image via a first decoder, and to the segmentation via a second decoder, and to the classification of COVID and non-COVID image via a multi-layer perceptron (see Fig 4).

**Fig. 4.**
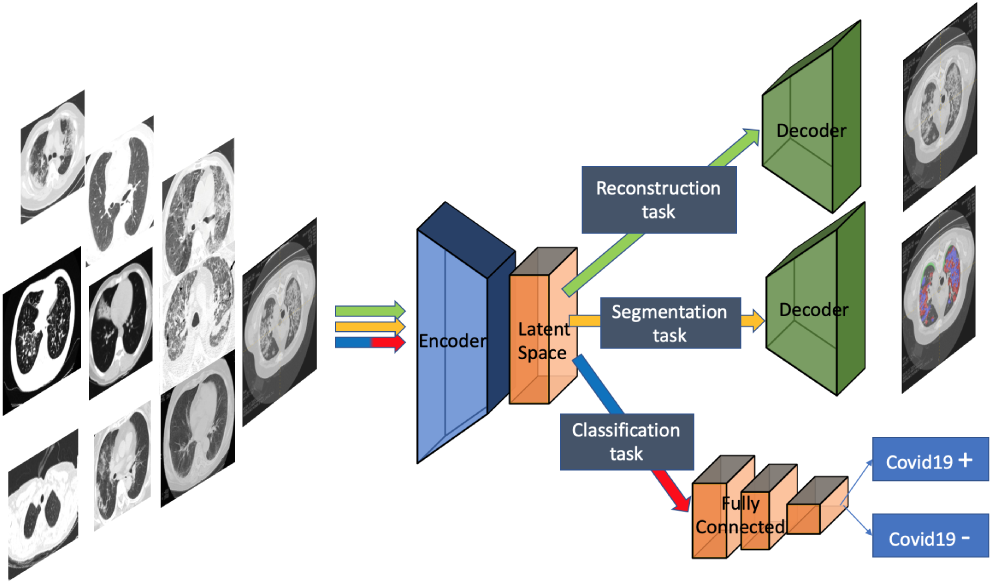
Our proposed architecture, composed of an encoder and two decoders for image reconstruction and infection segmentation. A fully connected layers are added for classification (COVID vs non-COVID)

#### 2.2.1. The Encoder-Decoder

the encoder-decoder is a 2D U-NET for both reconstruction and segmentation tasks. The encoder is a 10 convolutional layers with features maps from 64 to 1024 while the decoder is 10 layers of convolutions with upsamlping and 4 additional convolutional layers.

#### 2.2.2. The reconstruction task T1

We trained the model with a linear activation for the output and a mean squared error for the loss function (loss1) and with accuracy as the metric:

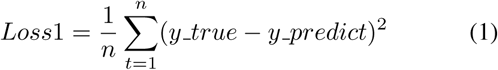

#### 2.2.3. The segmentation task T2

we used the same architecture as the reconstruction except for the activation function for the output, which is a sigmoid. The loss function is based on the dice coefficient loss (loss2) which is considered as the metric:

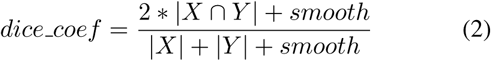

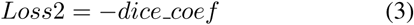

where the smoothing factor to avoid a division by zero.

#### 2.2.4. The classification task T3

the output of the encoder is a tensor of mini batch × 32 × 32 × 1024 to which we add a convolutional layer followed by a maxpooling, and then a flatten operation to convert the data to a mono-dimensional tensor to perform the classification. The multi-layer perceptron consist of a two Dense layer with 128 and 64 neurons respectively, with a dropout of 0.5 and the activation function *elu*. The last layer is a Dense layer with one neuron for image classification using a *sigmoid* activation and a binary cross entropy as the loss function (loss3):

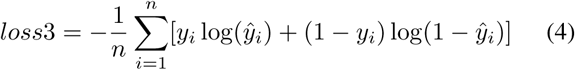

which is a special case of the multinomial cross-entropy loss function for m = 2 :

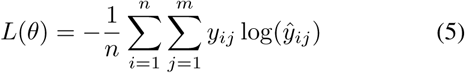

where n is the number of patients, y is the COVID label (binary, 1 if the patient has COVID, 0 otherwise) and 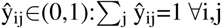 is the prediction of a COVID presence. In our experiments, the Adam optimizer [27] algorithm was used with a mini batches of 4 and a learning rate of 0.0001 for the 3 tasks to optimize the global loss function(loss glob):

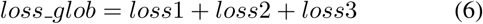

Our model was trained for 500 epochs with an early stopping of 35.

### 2.3. Implementation

The implementation of our method was done using the keras library with tensorflow in backend. The model was performed on an nvidia p6000 quadro gpu with 24gb, and 128 RAM.

## 3. EXPERIMENTATION

We conducted three experiments to evaluate our model.

### Experiment 1

The first experiment consisted of tuning the hyperparameters and add/remove a task to find the best model using only the training dataset. Several models were developed by combining the tasks 2 by 2 and the 3 tasks with different resolutions of images (512 × 512 and 256 × 256). The combination of the first task and the second one is only to evaluate segmentation results, since it is for image reconstruction and infection segmentation, while the peer T1 and T3 is for classification.

### Experiment 2

The second experiment consisted of comparing our model with state of the art method U-NET to compare the performance on the segmentation task. Two U-NET with different resolutions were trained: 512 × 512 and 256 × 256. In Fig 6 a comparison between our model and U-NET for infection segmentation.

**Fig. 5.**
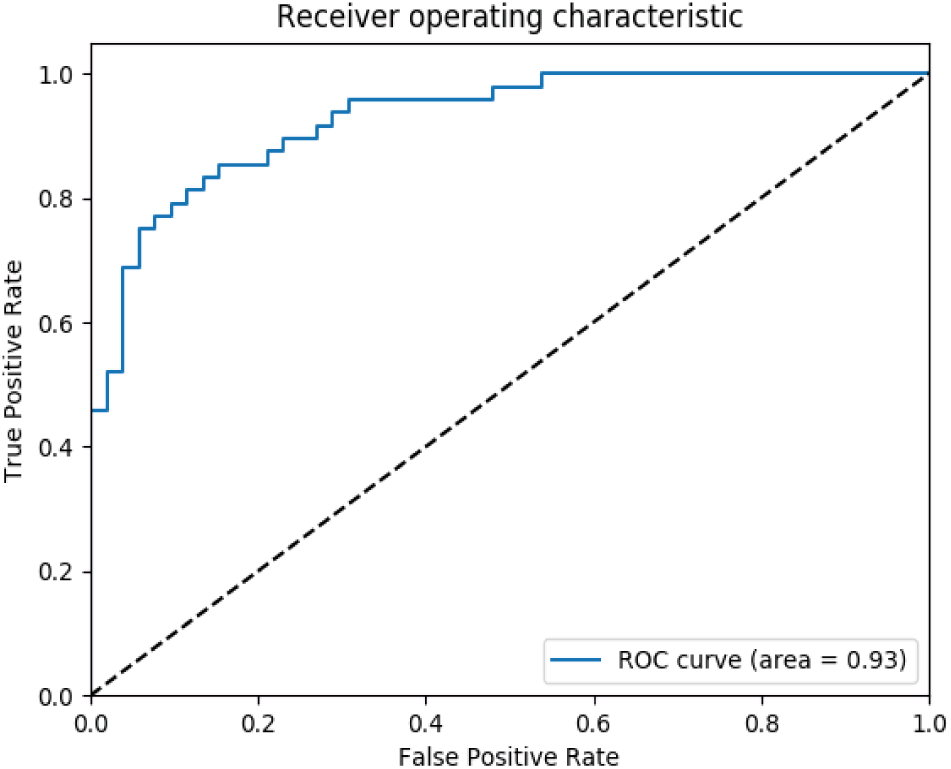
ROC curve of our best model, with an area under the curve (AUC)= 0.93.

**Fig. 6.**
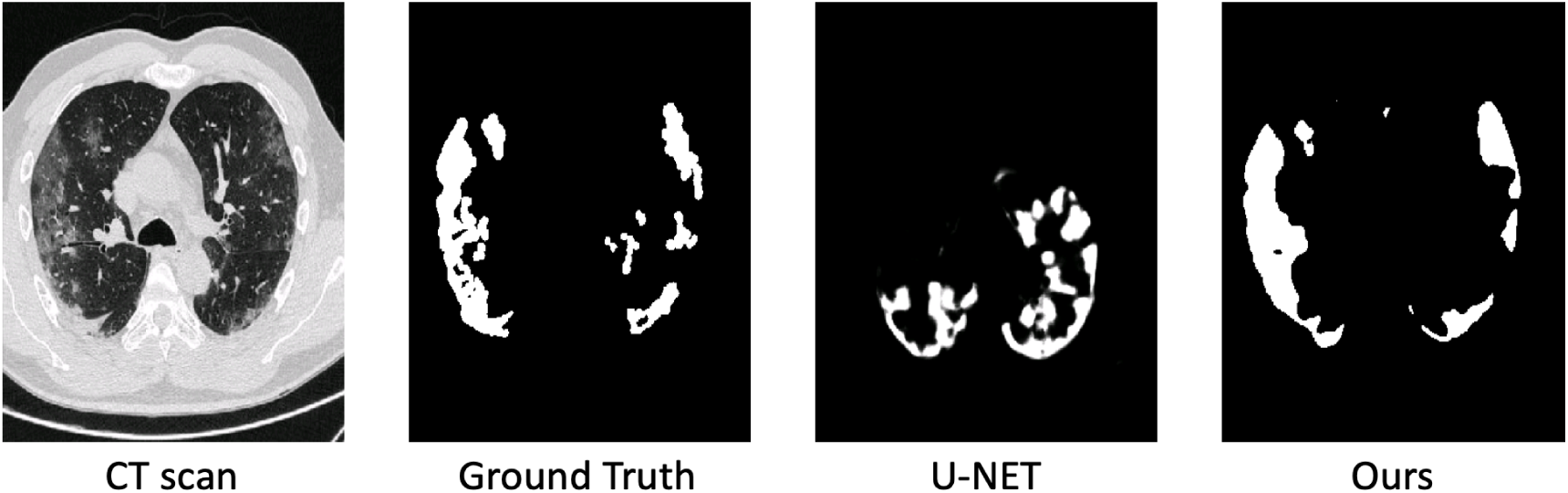
A comparison between our model and U-NET for infection segmentation.

### Experiment 3

The third experiment was the comparison between our model and convolutional neural network trained to perform classification only. The CNN used is an 8 layer deep neural network with 6 convolutional layers, where each one is followed by a Maxpooling and a Dropout regularization of 25% to prevent the model from overfitting. The feature maps go from 8 to 256 by a factor of 2 between each two layers. We used 3 × 3 filter for convolution and 2 × 2 for Maxpooling. Then a Flatten followed by two Dense layers with 128 neurons and 1 neuron respectively. A Dropout of 50% is also applied to the first layer to reduce and prevent overfitting. The activation function is *elu* for all layers except the last one which is a *sigmoid* to generate a probability for each class COVID vs non-COVID. The loss function is the binary cross-entropy and the metric is the accuracy, with the Adam optimizer. The CNN was optimized in order to ensure a fair comparison with our proposed model. The model was trained for 1500 epochs with an early stopping of 35, in the same condition as our model.

## 4. VALIDATION METHODOLOGY

For the validating methodology, we split the data for training, validation and test as shown in Table 1. Among the 349 COVID cases in the training, the ground truth for the infection label (segmentation task) was available for 50 CT scans. Twenty others were in the validation and thirty in the test set. For non-COVID cases, different kinds of pathology such as lung cancer, pneumonia or normal cases were selected randomly in train, validation and test. For a fair comparison, the other methods were trained, validated and tested in the same group of data. The performance of the models were evaluated using the dice coefficient for the segmentation task, and the accuracy (Acc), sensitivity (Sens), specificity (Spec) and area under the ROC curve (AUC) for the classification.

## 5. RESULTS

The main results of the three experiments are shown in Table 2. Metrics include: dice coefficient, accuracy, sensibility, specificity and the area under the ROC curve.

**Table 2.** Classification and Segmentation results: Experiment 1 for optimizing hyperparameters and choosing the best combination of tasks. Experiment 2 for segmentation analysis and Experiment 3 for classification.

|  | Method | Dice_coef | Acc | Sens | Spec | AUC |
| --- | --- | --- | --- | --- | --- | --- |
| Experiment 1 |  |  |  |  |  |  |
|  | T1 & T2 | 68.94% | / | / | / | / |
|  | T1 & T3 | / | 0.76 | 0.60 | 0.90 | 0.86 |
|  | T2 & T3 | 70.28% | 0.77 | 0.67 | 0.87 | 0.85 |
|  | T1 & T2 & T3 with 512 x 512 | 76.09% | 0.85 | 0.75 | **0.94** | 0.92 |
|  | **Ours (T1&T2&T3 256 x 256)** | **78.52%** | **0.86** | **0.94** | 0.79 | **0.93** |
| Experiment 2 |  |  |  |  |  |  |
|  | **Ours** | **78.52%** | **0.86** | **0.94** | **0.79** | **0.93** |
|  | U-NET 512 x 512 | 67.14 % | / | / | / | / |
|  | U-NET 256 x 256 | 69.09 % | / | / | / | / |
| Experiment 3 |  |  |  |  |  |  |
|  | **Ours** | **78.52%** | **0.86** | **0.94** | **0.79** | **0.93** |
|  | CNN 8-layers | / | 0.73 | 0.75 | 0.71 | 0.83 |

### Experiment 1

As shown in Table 2, the best dice coefficient = 78.52%, accuracy (acc = 0.86) and area under the curve (auc = 0.93) were obtained with the combination of the three tasks of image reconstruction, infection segmentation and image classification, with all images resized to 256 × 256. The results of 4 other experiments were also shown with multi-task learning but with a higher resolution of 512 × 512, and the combination 2 by 2 of the other tasks. The major differences between our best model and the model with higher resolution are in the sensitivity (0.94 vs 0.75) and specificity (0.79 vs 0.94). Compared to the peer combination of T1 and T3 for segmentation our model proved to be more performing with an improvement of +7% of dice, and a higher AUC and accuracy compared ti the peer T1 & T3 for classification only. The same result was observed for the peer segmentation & classification without reconstruction. Those results confirm the usefulness of the use of the reconstruction task to extract meaningful features and help improve the results of the other two tasks. The ROC curve of our model is shown in Fig 5.

### Experiment 2

In Table 2, the best result for image segmentation was obtained using our method with a dice coef of 78.52% versus 69.09% and 67.14% using U-NET with 256 × 256 and 512 × 512 resolutions respectively. The combination of the reconstruction, segmentation and classification results in a higher accuracy to detect infection regions, compared to the use of the U-NET model alone.

### Experiment 3

The results of experiment 3 also are given in Table 2. We compared our multi-task deep learning model with a deep convolutional neural network. The obtained results show that our model outperformed the CNN in both accuracy and AUC.

## 6. DISCUSSION

We have developed a new deep learning multi-task model to jointly detect COVID-19 CT images and segment the regions of infection. We have also evaluated several the state of the art algorithms such as U-NET and CNNs. To obtain our best model, we tested the different combinations of tasks 2 by 2 and all the 3 tasks simultaneously with different images resolutions. Our motivation was to leverage useful information contained in multiple related tasks to help improve both segmentation and classification performances.

In addition to the many advantages of using CT images to spot early COVID-19 patients and isolate them, deep learning methods using CT images can be used as a tool to assist physicians fighting this new spreading disease, as they can be used also to not only classify and segment images in the medical field, but also to predict the outcome of treatment for example [28]. Other powerful tool which come to hand is the use of weakly supervised learning algorithms to classify the image and detect the lesions where only a few datasets are available [29]. These weakly supervied methods can help the progress in the fight against the coronavirus COVID-19 where only few databases are usually available, and physicians are not able to provide many labeled data.

## 7. CONCLUSION

In this paper, we proposed a multi-task learning approach to detect COVID-19 from CT images and segment the regions of interest simultaneously. Our method can improve the segmentation results even if we do no have many segmentation ground truths, thanks to the classification data with ground truth which can be easily obtained compared to that of segmentation. Our method shows very promising results. It outperformed the state of the art methods for image segmentation when used alone such as U-NET or image classification such as CNNs. We have shown that by combining jointly these two tasks, the method improves for both segmentation and classification performances. Moreover, adding a third task such as image reconstruction, the encoder can extract meaningful feature representation which can help the other tasks (classification and segmentation) to improve even more their performances.

We have shown also that we can obtain very good sensitivity from CT images, which can tackle the need to detect infected people at an early stage to isolate them, and therefore, to limit the spreading of the disease. In future work, we will test our method on a larger database to confirm its good performance.

## Data Availability

Public data and data from Henri Becquerel Institution were used in this work

## Notes

### Competing Interest Statement

The authors have declared no competing interest.

### Funding Statement

None

